# Models of Accommodation Deficiency in Presbyopia Patients

**DOI:** 10.1101/2020.08.12.20173724

**Authors:** Mohit U. Karkhanis, Aishwaryadev Banerjee, Chayanjit Ghosh, Rugved Likhite, David Meyer, Carlos H. Mastrangelo

## Abstract

**Purpose:** We describe new phenomenological illumination-dependent static models of the accommodation deficiency for patients with presbyopia. Such models are suitable for vision restoration with adaptive-optics accommodating eyeglasses and contact lenses.

**Methods:** Data from fifteen participants over the age of 45 and diagnosed with presbyopia was collected. Participants were asked to wear a pair of mechanically-tunable eyeglasses and clearly identify the optotypes corresponding to the LogMAR 0.0 line on Early Treatment of Diabetic Retinopathy Study (ETDRS) charts, by suitably adjusting the optical powers of the lenses on these tunable eyeglasses for each measurement. Seven ETDRS charts, placed at distances from the patients varying from 4 m through 30 cm, were used under three chart illumination levels (75 lx, 500 lx and 800 lx). The optical power of the lenses in the patient-adjusted tunable eyeglasses was subsequently measured using a Shack-Hartman wavefront sensor for each chart, which provided the accommodation deficiency data of the participants.

**Results:** The measured accommodation deficiency data from 15 presbyopes was curve-fitted to a model for each patient. The calculated root-mean-square error values for the fitted models ranged between 0.09 D – 0.67 D over a 3.08D accommodation stimulus range.

**Conclusions:** The data shows that while accommodation deficiency in humans is a function of the stimulus, it is also strongly dependent on the object illumination and age of the patients. The models adequately describe the relation between static accommodation deficiency, accommodation stimulus and object illumination.

## Introduction

A healthy human eye has the ability to accommodate vision to near and far targets. In the first two decades of life, accommodative amplitude of the human eye has been shown to remain relatively stable in the range of 7-10 D. Long term studies suggest this accommodative amplitude reduces almost linearly with progressing age and reaches to 0.5-1.5 D by the age of 50, ultimately causing visual impairment^1-3^ and presbyopia^4^. This continuous age-related irreversible accommodation degradation is specified by Duane’s loss curve^4–7^.

Currently presbyopia is treated utilizing multifocal and progressive corrective eyewear. The major difficulty with these approaches is that they severely reduce the in-focus field of view and in fact do not restore normal vision. Fundamental restoration of normal vision requires lenses that can change optical power based on the object distance as a normal human crystalline lens does. Recent advances in adaptive optical systems^8^ suggest that such smart eyewear systems can be implemented in lightweight form for eyeglasses and more recently, contact lenses configurations. These advances in adjustable-focus lens technologies and low-power electronic systems have facilitated the development of accommodation-correction devices like smart-eyeglasses, which can provide automatic accommodation and potentially restore near-normal vision in presbyopes^9–12^. Smart-eyeglasses consist of three main electronic components – sensors, embedded computing systems and adjustable focus lenses. They use a host of sensors to determine the object distance^9,10,12^ and the gaze (point of visual focus) of every eye^11,13^. Using this data, the embedded computing systems within these devices can near-instantaneously change the optical power of the adjustable-focus lenses by performing quick calculations which use real-time sensor data and electronically-stored accommodation data of the patient. A recent study done in 2019 showed that the visual acuity of presbyopes improved while wearing a novel accommodation-correction device^11^. The participants in that study also preferred wearing the accommodation-correction device over their regular prescription eyeglasses. These devices hold great promise as they can restore normal vison and improve the lives of billions of people affected by presbyopia.

An essential requirement for full accommodation restoration is detailed knowledge of the individual’s accommodation deficiency (AD). AD is the difference between the accommodation stimulus (reciprocal of the object distance) and the individual’s accommodation response (AR) and describes the amount of defocus error present in the individual’s ocular system. The adaptive eyewear can fully restore normal vision by compensating for this deficiency and thus negating the effects of presbyopia. While this correction concept can in principle be performed for any presbyopia affected individual, the procedure requires detailed measurement of the AD curve which may not be always practically feasible. However, this procedure can be simplified through the use of semi-empirical models which can be realized with just a few AD measurements. At present, such models are poorly understood. The objective of this paper is the development of accurate models for the AD curve that can be fitted to the individual AD characteristics, subject to various illumination and object distance conditions with as few measurements as possible. In the sections below we first briefly survey the existing models for AD in the literature and then discuss an improved set of models which can fit the characteristics of a broad set of individuals. Experimentally, we measure the AD characteristics of a set of 15 individuals to construct these models and we discuss the impact on the accuracy of the models and the AD restoration with them in terms of the vision improvement realized with the model in terms of the effective individual visual acuity.

## Inadequacy of Current Accommodation Deficiency Models

A literature survey reveals a significant variation in the accommodation loss curve between individuals^4-7^. Moreover, every individual’s eye behavior is characterized by its accommodation response curve which maps the accommodation stimulus (reciprocal of the object distance) to the actual change in the optical power exhibited by the eye. In an ideal eye, the slope of the accommodation response curve should be one, but as presbyopia develops, the response flattens to lower optical powers^14,15^ as seen in Figure (1).

**Figure 1.**
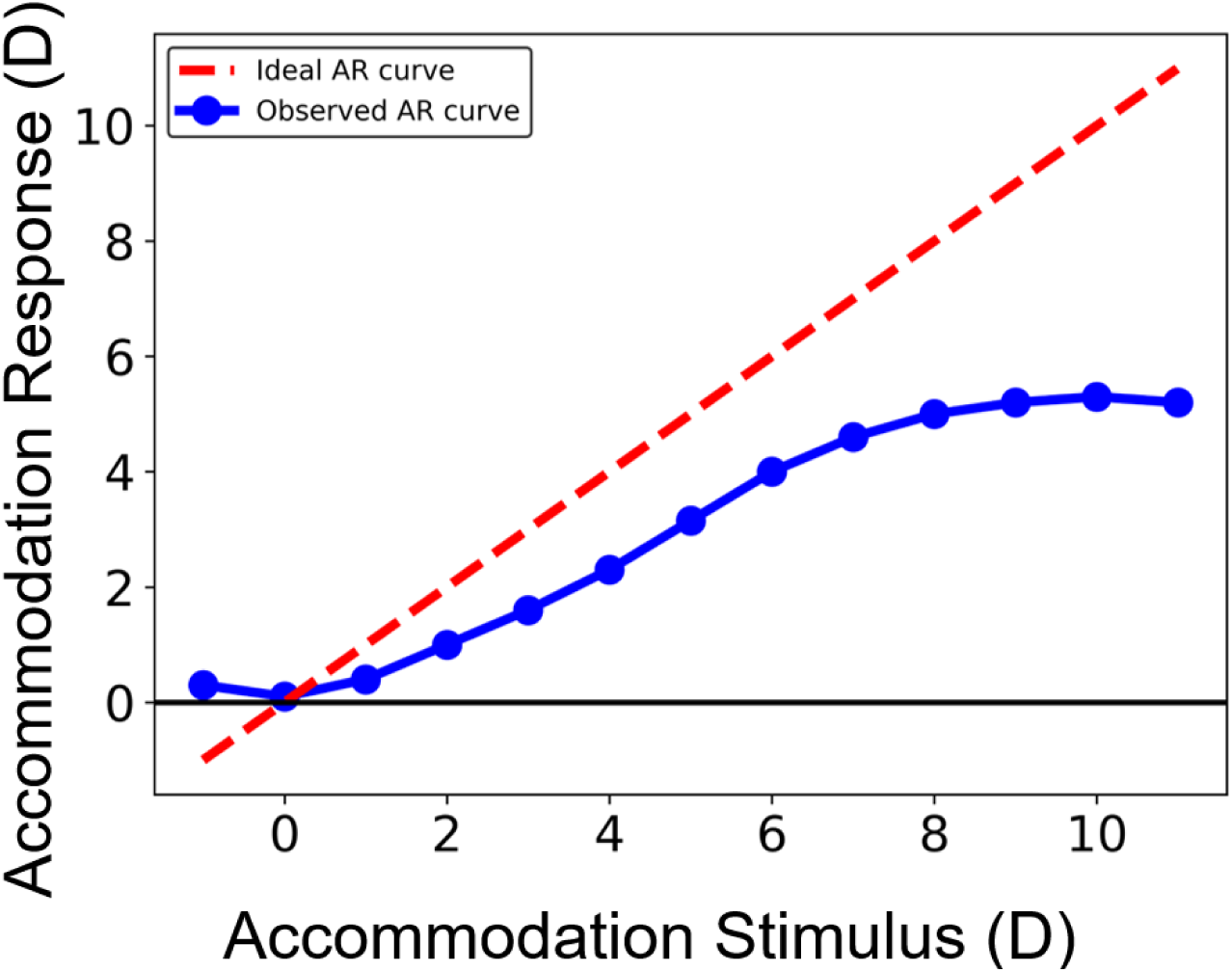
Example of typical *S-type* accommodation response curve for the human eye. The curve progressively flattens for higher powers with age^14,15^.

Two different phenomenological models for the accommodation characteristics of presbyopes have been suggested so far which originate from the two major variants of the lenticular theory^14–29^. In the Helmholtz-Hess-Gullstrand (HHG) theory, the loss in accommodation is completely attributed to the morphological changes in the lens and the lens capsule. In this model, a constant amount of ciliary muscle contraction is required throughout life to produce a unit change in accommodation and the ciliary muscle’s contractive force is independent of the patient age. The accommodation response corresponding to HHG theory is shown in Figure (2A). The model depicts a near-ideal response to stimulus over the manifest zone and a sharp transition into a hard saturation region or the latent zone. As the amplitude of accommodation reduces only due to lenticular changes with age, an increasing amount of ciliary muscle contraction will produce no accommodation change as reflected in the nonresponsive latent zone. In contrast, the Donders-Duane-Fincham (DDF) theory attributes loss in accommodation with age to changes in the bio-mechanical properties of ciliary muscle. In DDF, the amount of ciliary muscle contraction necessary to produce a unit change in accommodation progressively increases with age. The reduction in accommodation amplitude with age is due to the progressive weakening of the ciliary muscle while the abilities of the lens and lens capsule remain unchanged. Thus, contradictory to HHG, the amount of potential ciliary muscle force in reserve progressively declines with age. Figure (2B) shows the corresponding accommodation characteristics of DDF.

**Figure 2.**
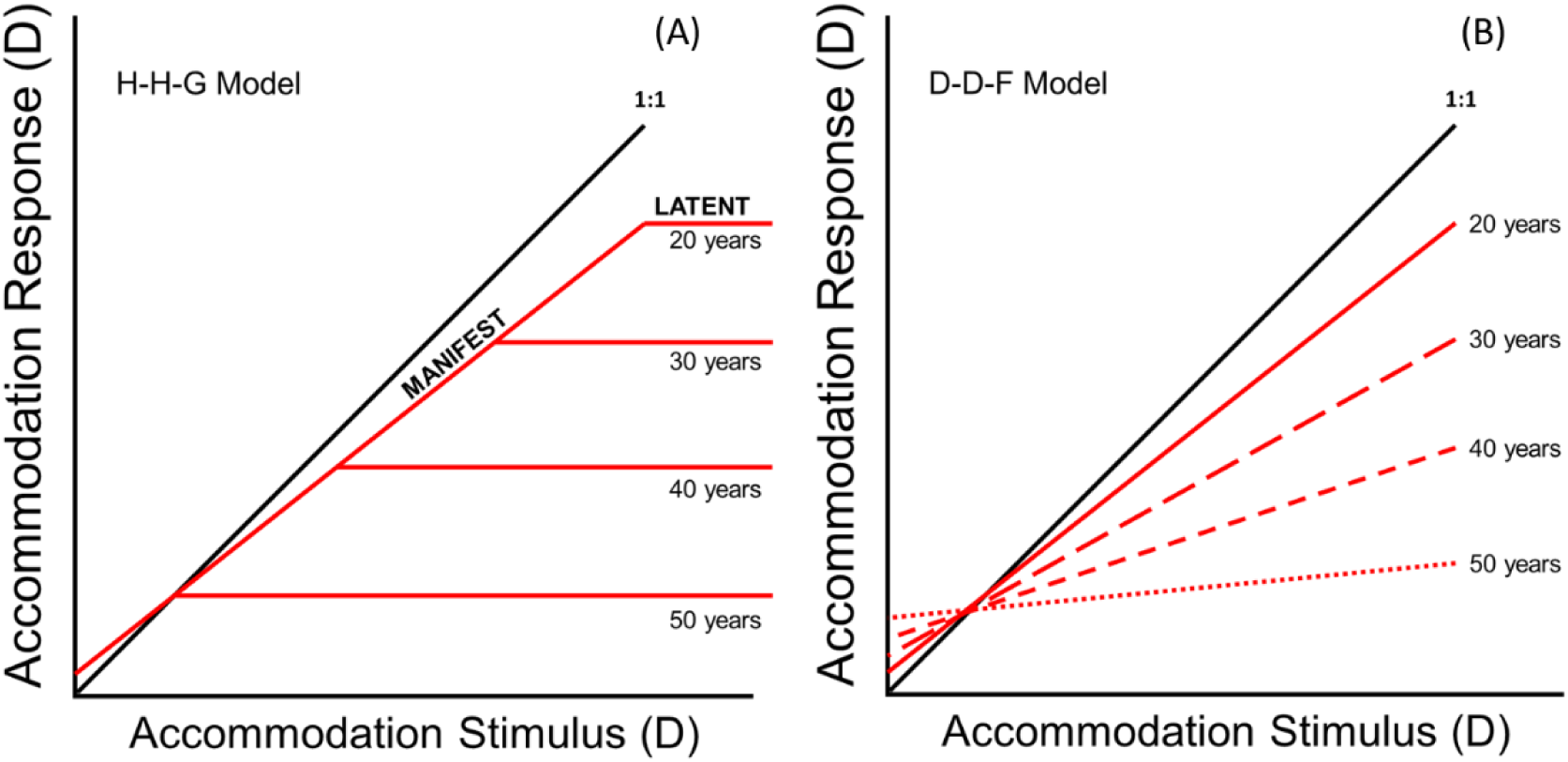
Comparison of accommodation response characteristics from the Helmholtz-Hess-Gullstrand and the Duane-Donders-Fincham models^14^. The black line is the ideal accommodation response. Neither model fits actual individual AD characteristics adequately for performing a good AD correction with adaptive smart corrective eyewear.

Neither of the HHG or the DDF models is accurate although the HHG model shows qualitative flattening of a typical accommodation response characteristics which resembles observations in presbyopes^15^. Furthermore, none of these established models include dependence on the ambient illumination level, which affects the dimensions of the pupil. With increasing illumination levels, the pupil size decreases^30^, thereby increasing the depth-of-focus of the visual system and reducing the required amplitude of accommodation to focus on an object. One of the principal reasons why presbyopes tend to squint is because doing so improves their visual acuity at the expense of much lower brightness. At lower illumination levels, the pupil size increases, reducing the depth of field and requiring more accommodation amplitude^31^. It has also been shown that accommodation depends on cone activity^32^. As the illumination levels are progressively lowered, the slope of the accommodation response curve steadily decreases. Eventually the accommodative system ceases to function (accommodation response curve becomes flat) when the cones are completely shut down and only the rods are active^33^. Such illumination dependent presbyopic behavior has not been studied previously.

Realization of practical and robust accommodation-correction devices requires extraction of accurate accommodation deficiency models from the presbyopic population that can represent the deficiency loss with errors < 0.5 D as in the average person, the RMS aberration of the eye is approximately 0.33 μm^34,35^ which, according to the relation given by Watson and Ahumada^36^, corresponds to a nearly 0.5 D error. Furthermore, the accommodation amplitude in most presbyopes is small but non-zero and it cannot be ignored as this response plays a major role in the final quality of image produced by accommodation correction devices. Therefore, to perform high-quality accommodation correction, it is necessary to know the detailed behavior of the accommodation deficiency in presbyopes under different object distances (stimulus) and illumination conditions and subsequently implement these findings into more accurate accommodation-correction devices.

## Material and Methods

### Human Study Participants

Human study approval was acquired from the University of Utah Institutional Review Board (IRB_00114415), and experiments were performed according to the ethical standards laid down in the Declaration of Helsinki, 1964. A total of 15 presbyopic subjects aged 45 – 68 years were recruited from a population of patients from the University of Utah Moran Eye Center and associated clinics. All subjects provided informed, signed consent before entry into the study. A record on clinical trials performed in this study has been registered with ClinicalTrials.gov^37^ (NCT03911596). Individuals with astigmatism >1.0 D, artificial intraocular lenses, or those having any ocular pathology that would inhibit accommodation of their natural lenses were excluded. The recruited subjects had eyeglass prescriptions between −1.5 D and +2.5 D and were correctable to 20/20.

### Study Procedure

Testing was carried out in a light proofed optometry exam room at the University of Utah Moran Eye Center. The visual task consisted of reading optotypes on 7 different ETDRS charts calibrated for 7 distances (4 m, 2 m, 1 m, 70 cm, 50 cm, 40 cm and 30 cm) under 3 chart illumination conditions (75 lx, 500 lx and 800 lx) using commercially available manually-tunable Adlens Hemisphere eyeglasses. Participants were assisted by the study staff in manually tuning the commercial tunable eyeglasses till they could correctly identify the optotypes corresponding to LogMAR 0.0 / 20/20 line on the ETDRS charts. Each lens in the eyeglasses was monocularly tuned before patients undertook the visual task binocularly under every test distance and illumination condition. To record the accommodation deficiency in participants, the optical power of the tuned lenses was measured every time using a Thorlabs WFS40 Shack-Hartmann wavefront sensor. Specially calibrated ETDRS charts, which were placed at distances of 1 m, 70 cm, 50 cm and 30 cm, were carefully designed and printed on high quality optical white paper. Chart retro-illumination was not used. Chart illumination was controlled using an LED studio lighting system which also provided diffused lighting for the exam room.

The correlated color temperature of the lighting system was fixed at 5000 K. Chart illumination level was kept constant while measuring the accommodation deficiency progressively from far (4 m) to near (30 cm) distances. Participants were tested under chart illumination levels of 500 lx, 800 lx and 75 lx, sequentially, and were given enough time to adjust their eyes before commencing the visual task. The study sessions lasted between 1.5 – 3 hours in duration.

### Unified Accommodation Deficiency Model

The typical experimental curve for presbyopic accommodation response as shown in Figure (1) displays a saturation type of response with flat regions at low and high accommodation stimuli and an approximately uniform slope in between. This type of accommodation response (AR) characteristic can be represented by the sigmoid shifted logistic function:

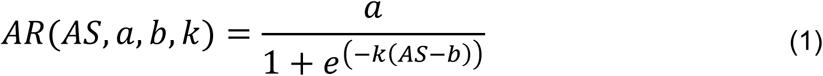

where *a, b* and *k* are 3 fitting parameters and variable *AS* represents the accommodative stimulus. The parameter *a* determines the accommodative amplitude of the individual i.e. the useful linear range of the experimental accommodation response curve. Parameter *k* determines range of stimuli for which the *AR* curve exhibits a linear response and parameter *b* corresponds to the midpoint of the non-flat accommodation stimulus range.

The accommodation response of presbyopes is a strong function of the illumination levels. Accommodation amplitude progressively reduces at lower lighting levels and ultimately approaches a fixed tonic value as shown by the experimental data in this article^33^. This degradation in the accommodation response is partially caused due to the increase in pupil diameter *(dp)* and consequently, the reduction in the depth of field at low illumination levels^31^. We approximate an initial candidate model for accommodation response as:

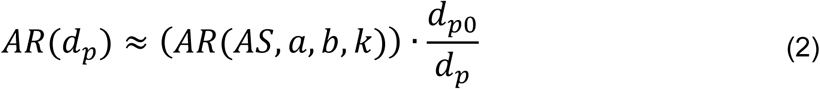

where *d_p0_* is the pupil diameter at a certain pre-decided reference light level and ratio *d_p0_/d_p_* is proportional to the ambient illumination levels. Equation (2) indicates that as pupil diameter decreases (high illumination levels), the accommodation response increases. This behavior is consistent with experimental evidence recorded by Ripps et al.^38^

Multiple clinical studies which describe the relation between pupil diameter and illumination level have been extensively reviewed by Watson and Yellot^39^. Out of the multiple models reviewed, the phenomenological De Groot-Gebhard model^40^ was found to be analytically simple yet precise in determining the relation between the pupil diameter and illumination levels. Using this model, the pupil diameter ratio in Equation (2) can be expressed as a function of illumination as:

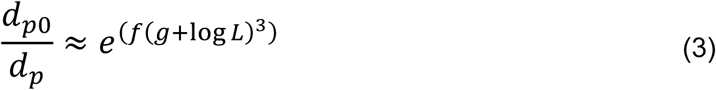

where *f* and *g* are dimensionless fitting parameters and *L* is the illumination level. Using Equations (1), (2) and (3), we can finally approximate accommodation response as:

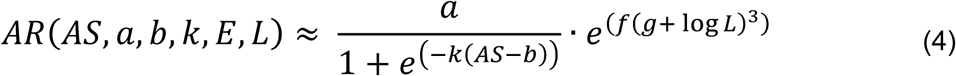

Since the accommodation deficiency *AD* = *AS* − *AR*, an illumination dependent accommodation deficiency model can be approximated as:

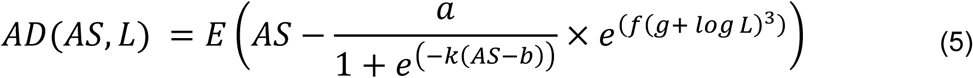

with *E* as an additional dimensionless fitting parameter. It can be seen in Equation (5) that the sigmoidal accommodation response improves with increasing levels of object illumination, consistent with phenomenological studies conducted by Johnson^33^ and Lara et al.^31^

## Results

A total of 15 subjects were recruited with mean (SD) age 54.6 (6.8) years. None of the participants exhibited astigmatism > 1.0 D, artificial intraocular lenses, or any other ocular pathology or surgery. Visual acuity of subjects was recorded in LogMAR after they completed the visual task using the manually-tunable adjustable-focus eyeglasses. Figures (3) – (5) show the corrected visual acuities of the subjects under varying accommodation stimuli and chart illumination levels. None of the 15 subjects exhibited corrected visual acuity worse than +0.2 LogMAR (20/32 Snellen equivalent). In order to better relate an individual’s visual acuity to eyeglass prescription, visual acuity scores corresponding to spherical refractive errors of 0.25 D and 0.5 D were calculated. To restore normal vision in presbyopes, accommodation-correction devices should provide vision correction with maximum spherical refractive error of 0.5 D which in practice, is in the same order as the spherical equivalent of the average RMS aberration of human eyes. According to the relation between spherical refractive error and visual acuity given by Smith^41^, the minimum angle of resolution *A* is related to the spherical refractive error *E* as

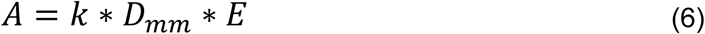

**Figure 3.**
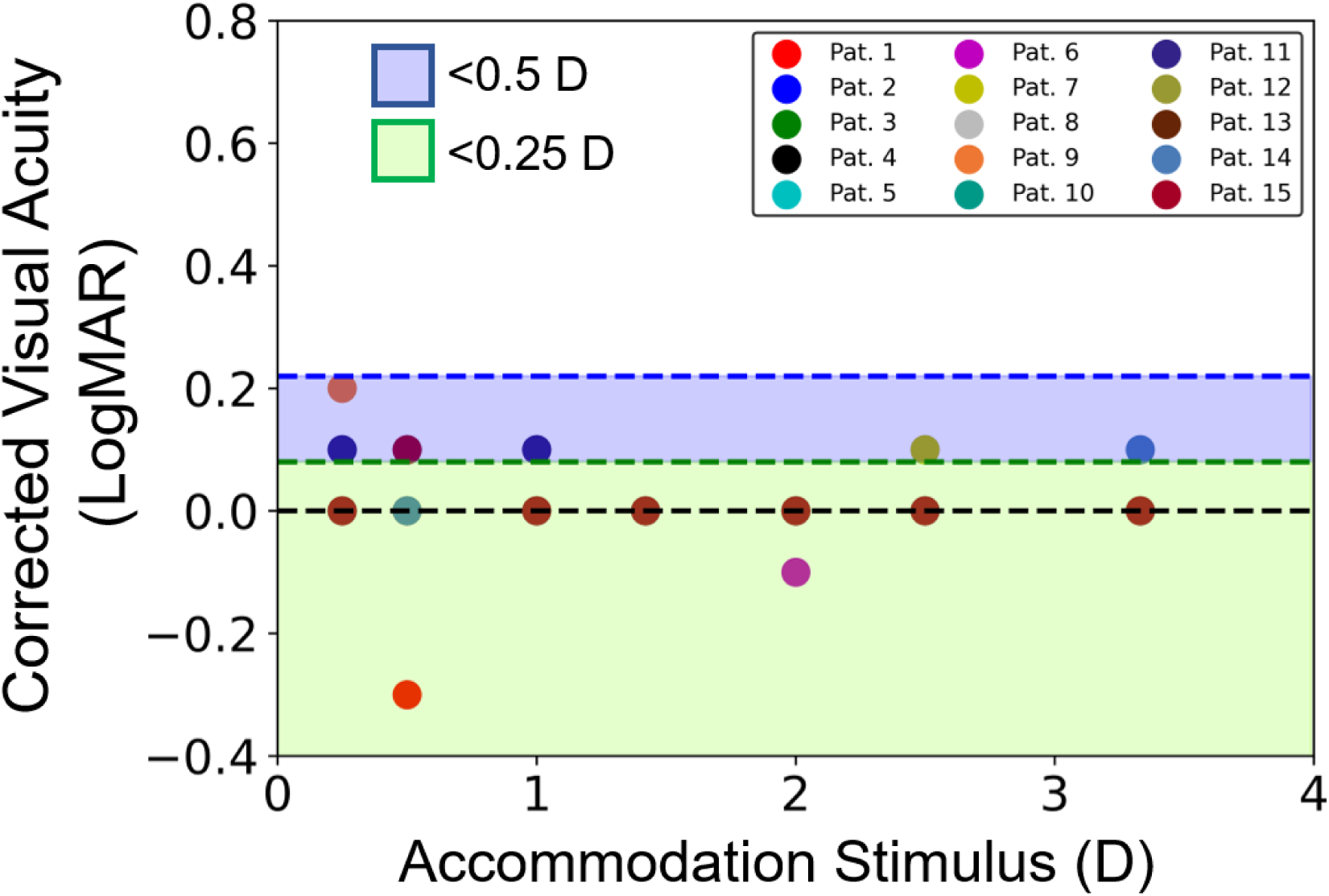
Corrected Visual acuities of 15 subjects at every accommodation stimulus under 75 lux chart illumination.

**Figure 4.**
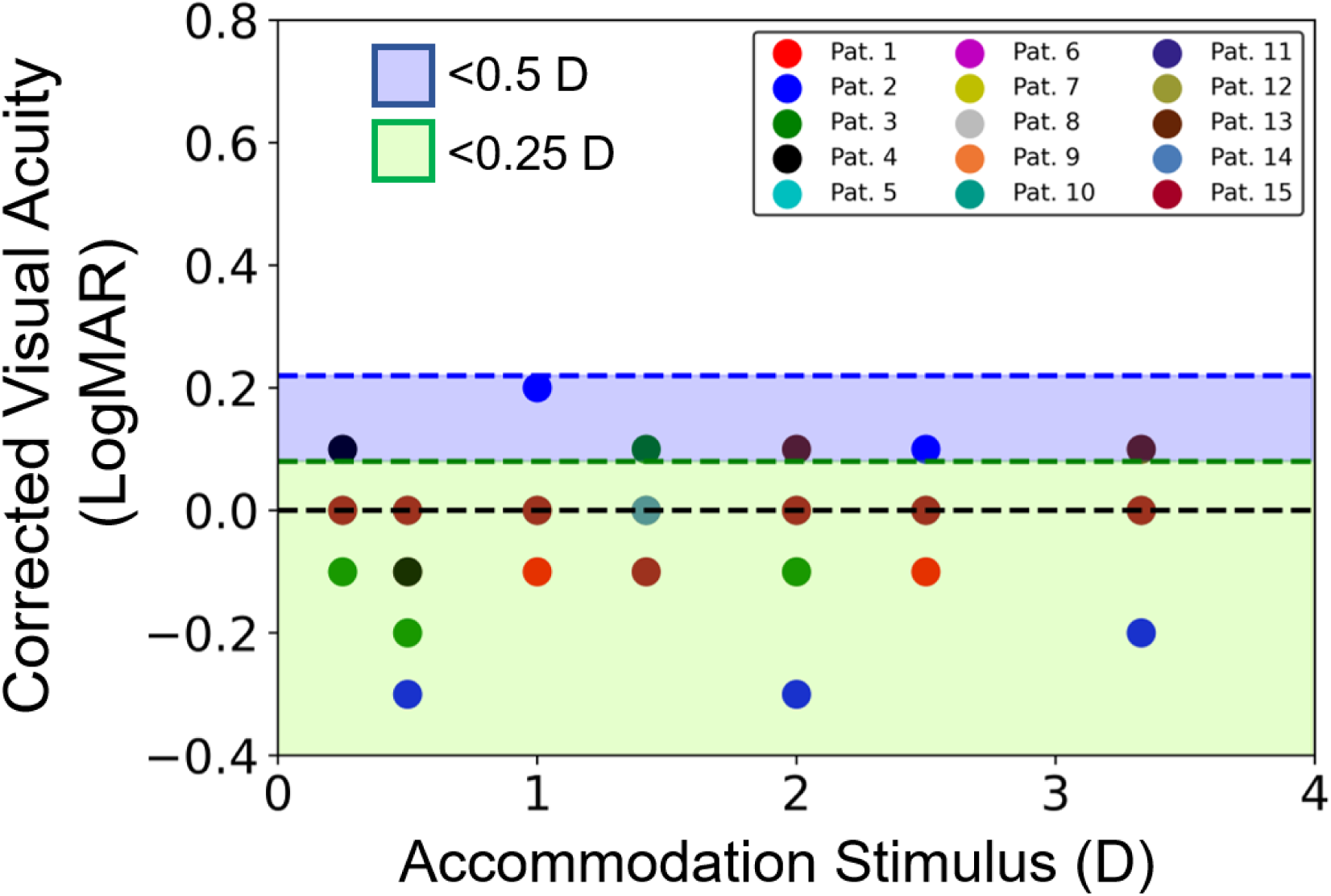
Corrected Visual acuities of 15 subjects at every accommodation stimulus under 500 lux chart illumination.

**Figure 5.**
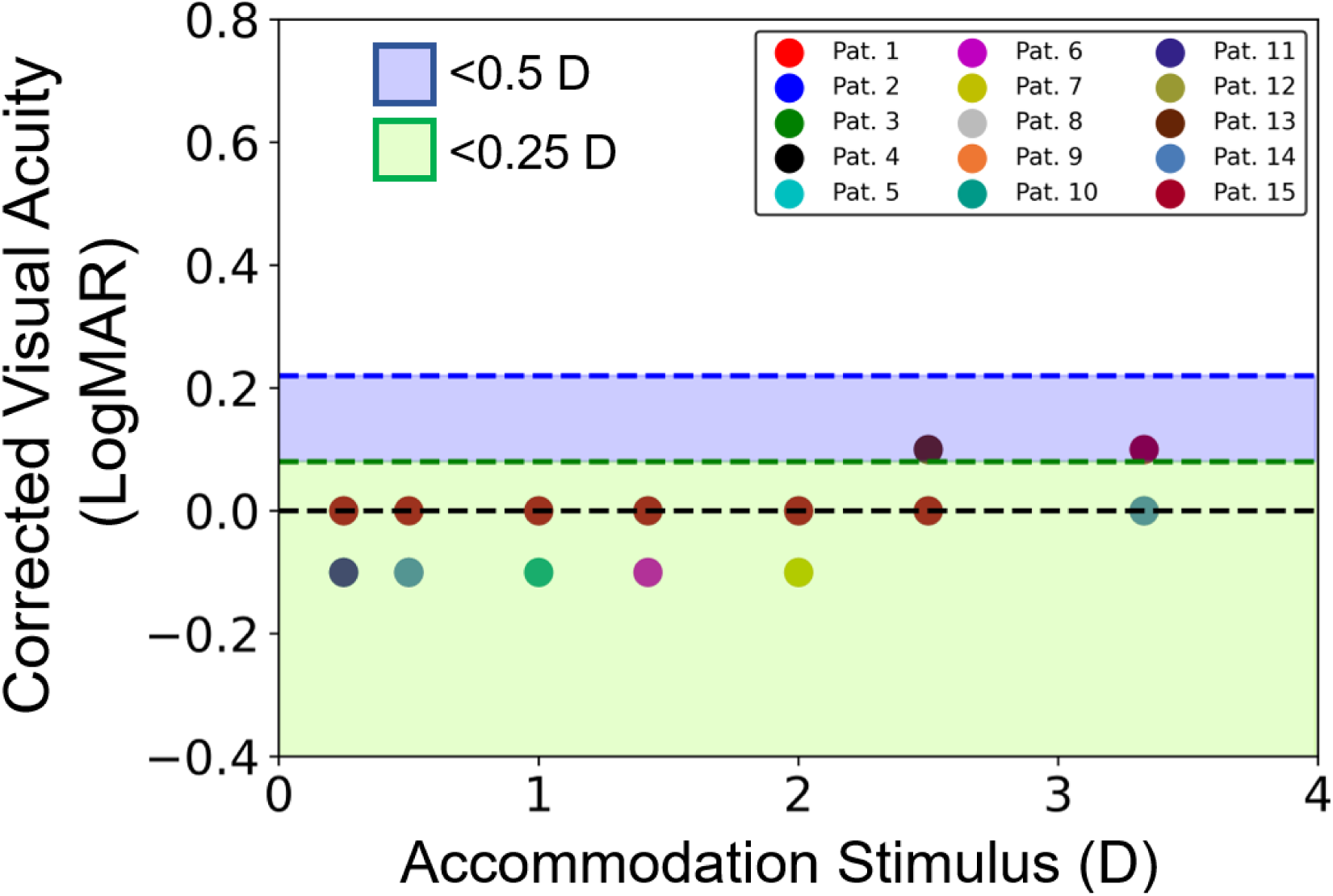
Corrected Visual acuities of 15 subjects at every accommodation stimulus under 800 lux chart illumination.

Where *D_mm_* is the pupil diameter in mm and *k* is a dimensionless constant with mean value of 0.83. Using Equation (6), for an average pupil diameter of 4 mm, spherical refractive errors of 0.5 D and 0.25 D yield visual acuity scores of +0.22 LogMAR and +0.08 LogMAR respectively. The blue and green lines in Figures (3) – (5) show these calculated visual acuity scores corresponding to spherical refractive errors of 0.5 D and 0.25 D, respectively. Corrected visual acuities falling within the blue region correspond to an acceptable spherical refractive error between 0.25 D and 0.5 D, while those falling within the green region correspond to a minimum desirable spherical refractive error of < 0.25 D in the accommodation-correction system.

Table (1) shows the recorded visual acuity of the subjects, averaged over all 7 distances, under the chart illumination levels of 75, 500 and 800 lux. A Wilcoxon signed-rank test^42^ was performed in MATLAB R2020a to determine the statistical significance of the changes in the observed corrected visual acuities under 75 lux and 800 lux. The improvement in the corrected visual acuities of subjects under a higher chart illumination level of 800 lux compared to 75 lux was found to be statistically significant (*p* = 0.0122, *α* = 0.05).

**Table 1.** Average corrected visual acuities of 15 patients under 3 chart illumination levels.

| Corrected VA (75 lux) | Corrected VA (500 lux) | Corrected VA (800 lux) |
| --- | --- | --- |
| -0.04 | -0.05 | -0.01 |
| 0.01 | -0.07 | 0.01 |
| 0.01 | -0.04 | -0.07 |
| 0.01 | 0.0 | -0.02 |
| 0.01 | 0.0 | -0.02 |
| 0.02 | 0.0 | -0.04 |
| 0.02 | 0.0 | -0.01 |
| 0.01 | 0.0 | 0.0 |
| 0.04 | 0.0 | 0.0 |
| 0.02 | 0.0 | -0.04 |
| 0.04 | 0.0 | -0.02 |
| 0.0 | 0.0 | 0.0 |
| 0.0 | 0.02 | 0.02 |
| 0.01 | 0.0 | -0.01 |
| 0.01 | -0.01 | 0.01 |

To study the accommodation deficiency characteristics in presbyopes, the observed accommodation deficiencies or defocus errors of the subjects were plotted against the accommodation stimulus (reciprocal of the chart distance). Figures (6), (7) and (8) show the observed accommodation deficiencies under chart illumination levels of 75 lux, 500 lux and 800 lux, respectively. It was found that the accommodation deficiency kept increasing with increasing accommodative effort, under all 3 illumination conditions.

**Figure 6.**
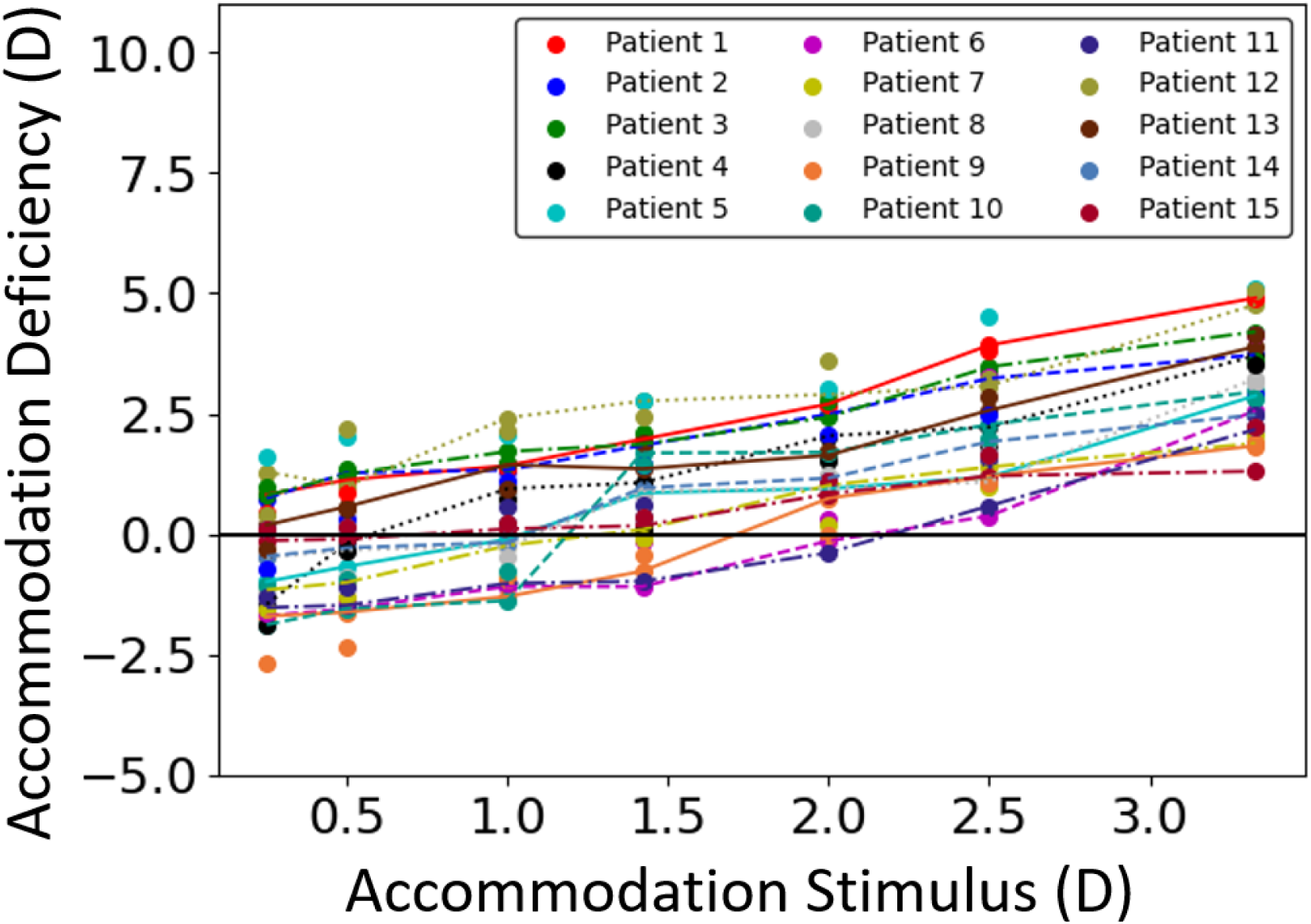
Measured accommodation deficiencies for 15 patients (30 eyes) under 75 lux chart illumination.

**Figure 7.**
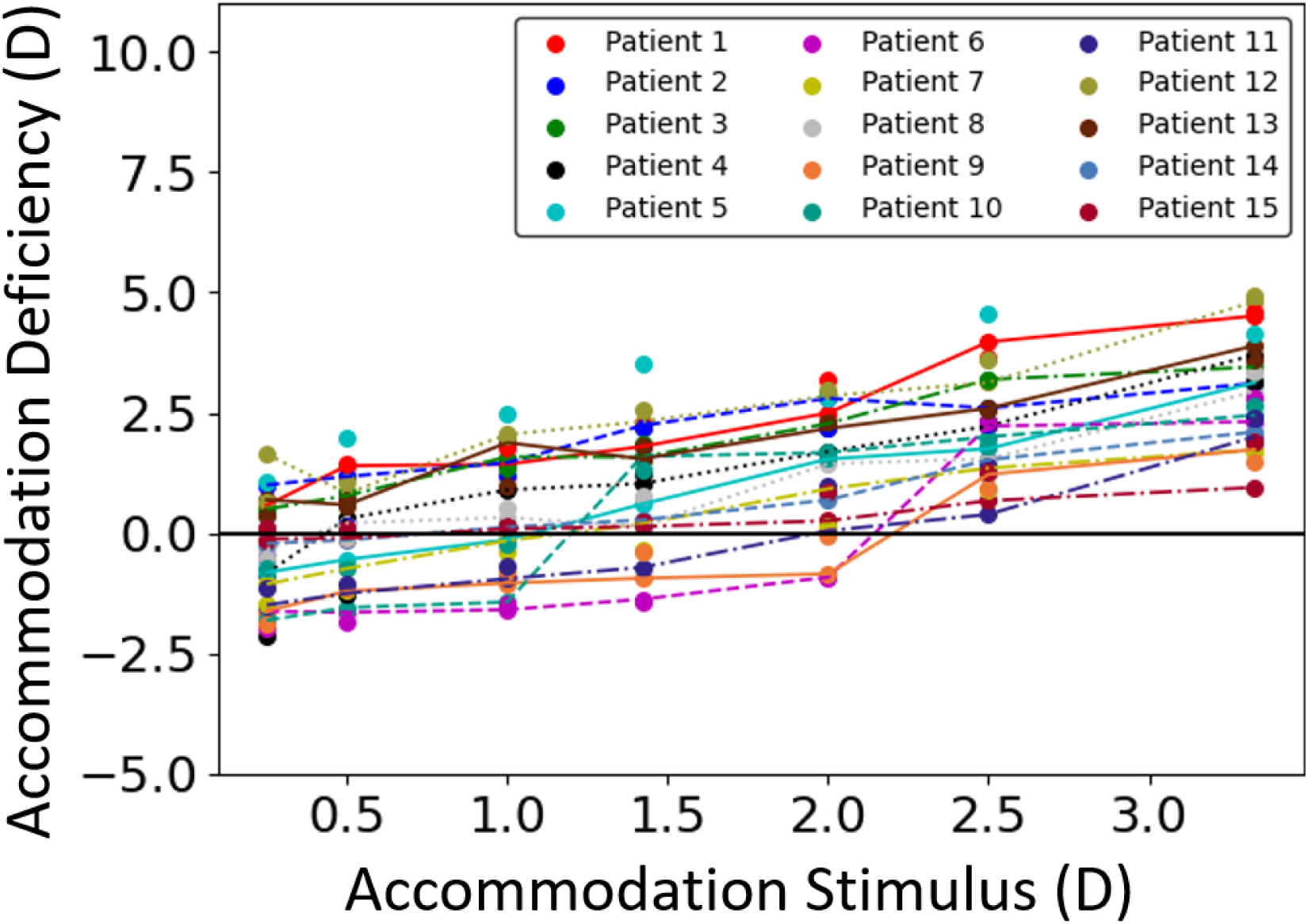
Measured accommodation deficiencies for 15 patients (30 eyes) under 500 lux chart illumination.

**Figure 8.**
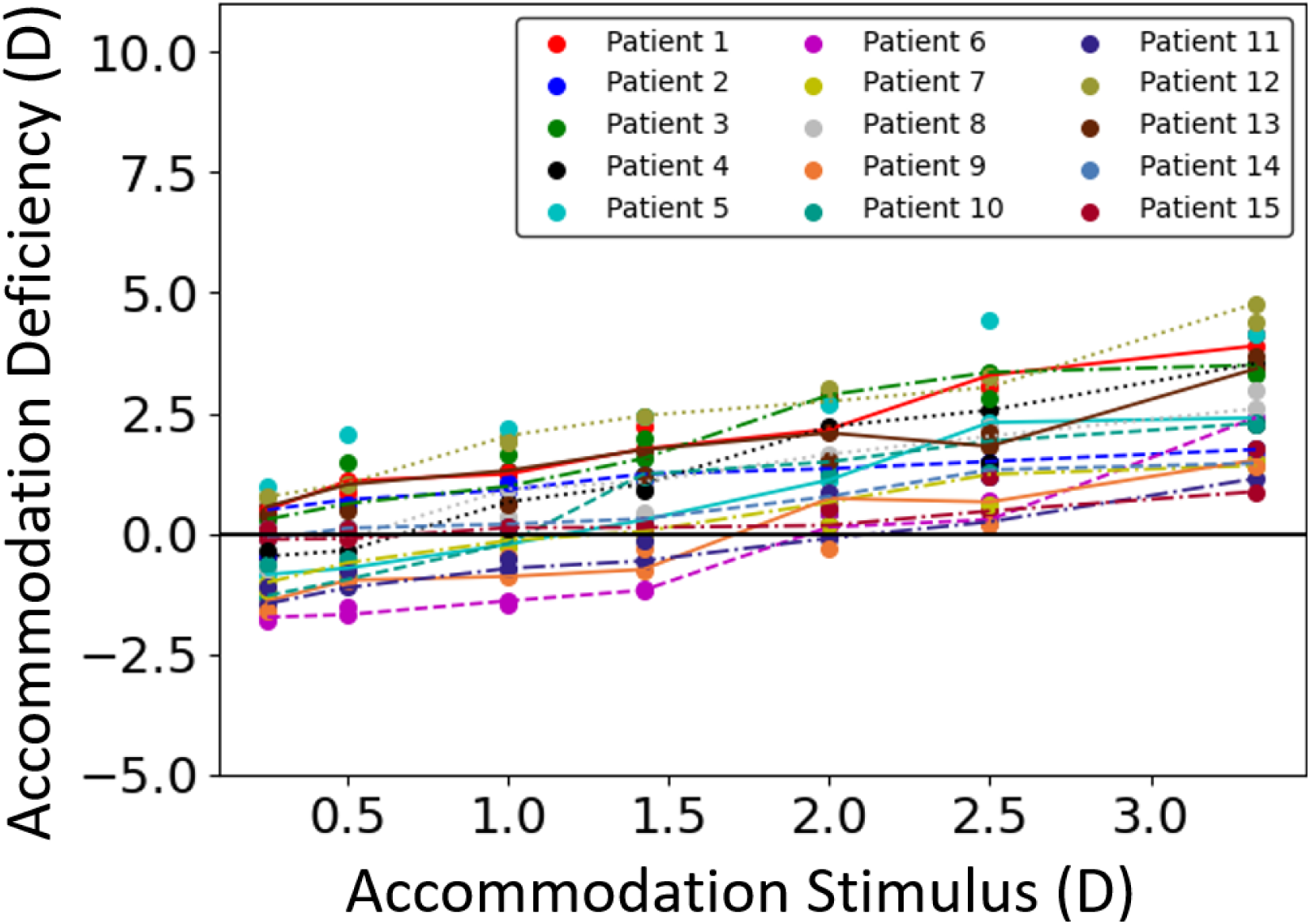
Measured accommodation deficiencies for 15 patients (30 eyes) under 800 lux chart illumination

Accommodation deficiency amplitude, which refers to the total change in the accommodation deficiency for an applied stimulus range, was also studied for every patient. Figure (9) shows the deficiency amplitudes for 15 subjects’ left and right eyes under different illumination levels. It also shows that for few patient’s eyes, the deficiency amplitude at 800 lux was greater than that at 500 lux. This seems counter-intuitive as accommodation deficiency decreases under higher object illumination levels due to improved depth of focus. However, there are some clues to explain this discrepancy. As light enters the eye, part of its energy is scattered in the crystalline lens^43^ and gives rise to a phenomenon called disability glare^44,45^. Disability glare is a physiological glare which generally impairs vision, decreases visual acuity without causing any discomfort^46^ and decreases contrast sensitivity^47^. Due to anatomical changes in the visual system with age^48–51^, senior people were found to be more sensitive to glare^52–55^. This could possibly explain why some eyes in this study exhibit higher accommodation deficiency amplitudes at 800 lux chart illumination compared to those at 500 lux chart illumination.

**Figure 9.**
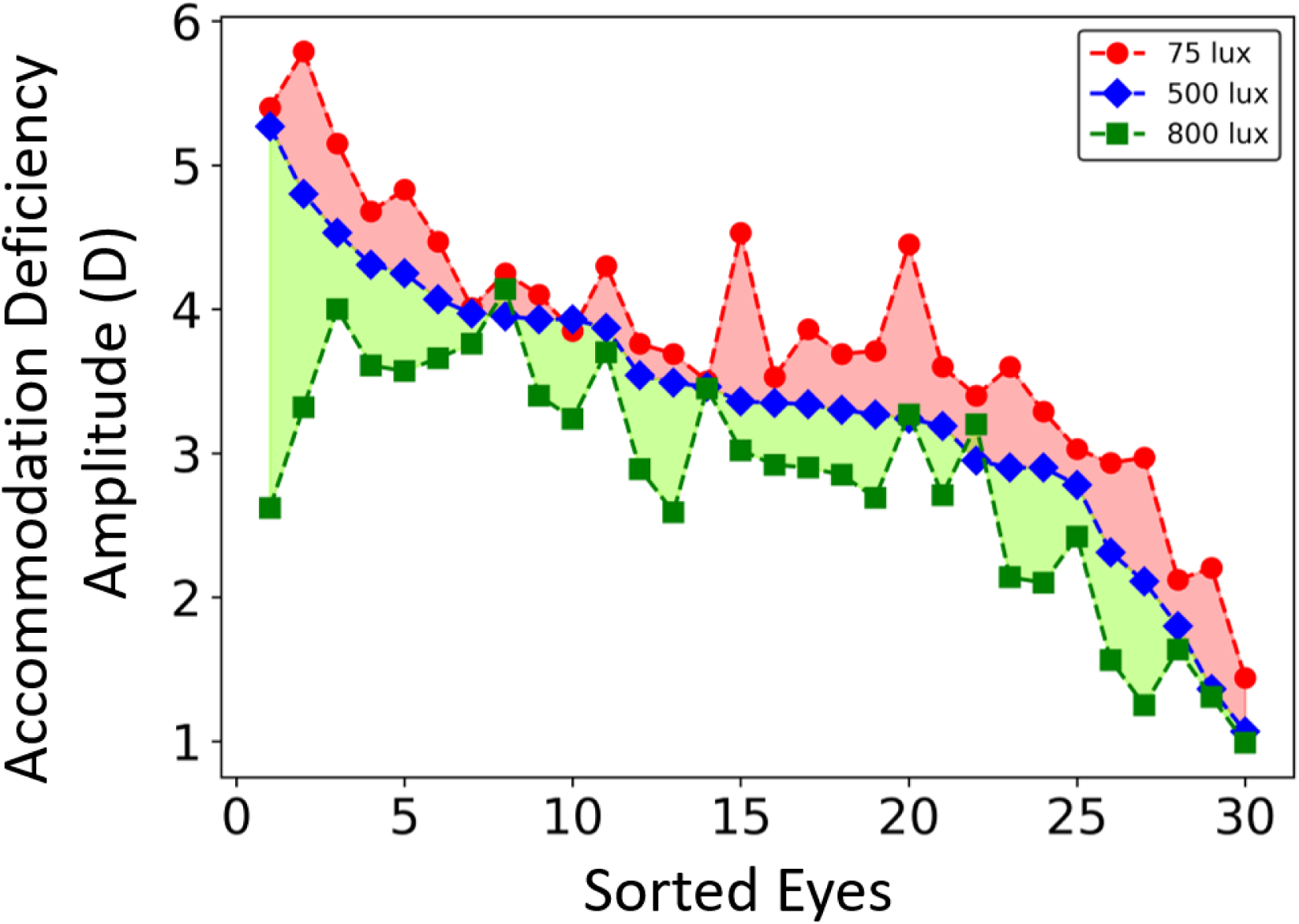
Observed accommodation deficiency amplitudes for 15 patients (30 eyes) under various illumination conditions. The eyes are sorted according to their deficiency amplitudes at 500 lux.

A Wilcoxon signed rank test^42^ was performed in MATLAB R2020a to determine the significance of the change in the accommodation deficiency amplitudes for both eyes under 2 different illumination levels of 75 lux and 800 lux. The deficiency amplitudes for both eyes were found to be lower for 800 lux chart illumination level compared to 75 lux chart illumination (left eyes: *p* < 0.001, *α* = 0.05, right eyes: *p* < 0.001, *α* = 0.05).

The observed accommodation deficiency data for each eye of all subjects were curve-fitted to Equation (1) utilizing the Levenberg-Marquardt nonlinear fitting algorithm^56,57^ in MATLAB R2020a.This yielded a separate characteristic equation and curve for every eye of every subject. Curve fits are usually analyzed with the R^2^ (goodness of fit / coefficient of determination) metric, however, this alone cannot determine whether the coefficient estimates are biased^58^ and we therefore require additional assessment of the residual plots after curve-fitting. A modified version of this parameter, called the adjusted-R^2^, is a parameter which has been adjusted for the number of curve-fitting coefficients^59^ and is always lower than the R^2^. Accordingly, to account for the number of coefficients in the presented model, the adjusted-R^2^ metric was used to analyze the curve fits. Figure (10) shows the monocular deficiency characteristics for 4 subjects under 500 lux chart illumination level recruited in the study. The adjusted-R^2^ (goodness of fit) values for 15 subjects ranged between 0.71 – 0.99 and the root-mean-square error was found to be between 0.03 D – 0.67 D.

**Figure 10.**
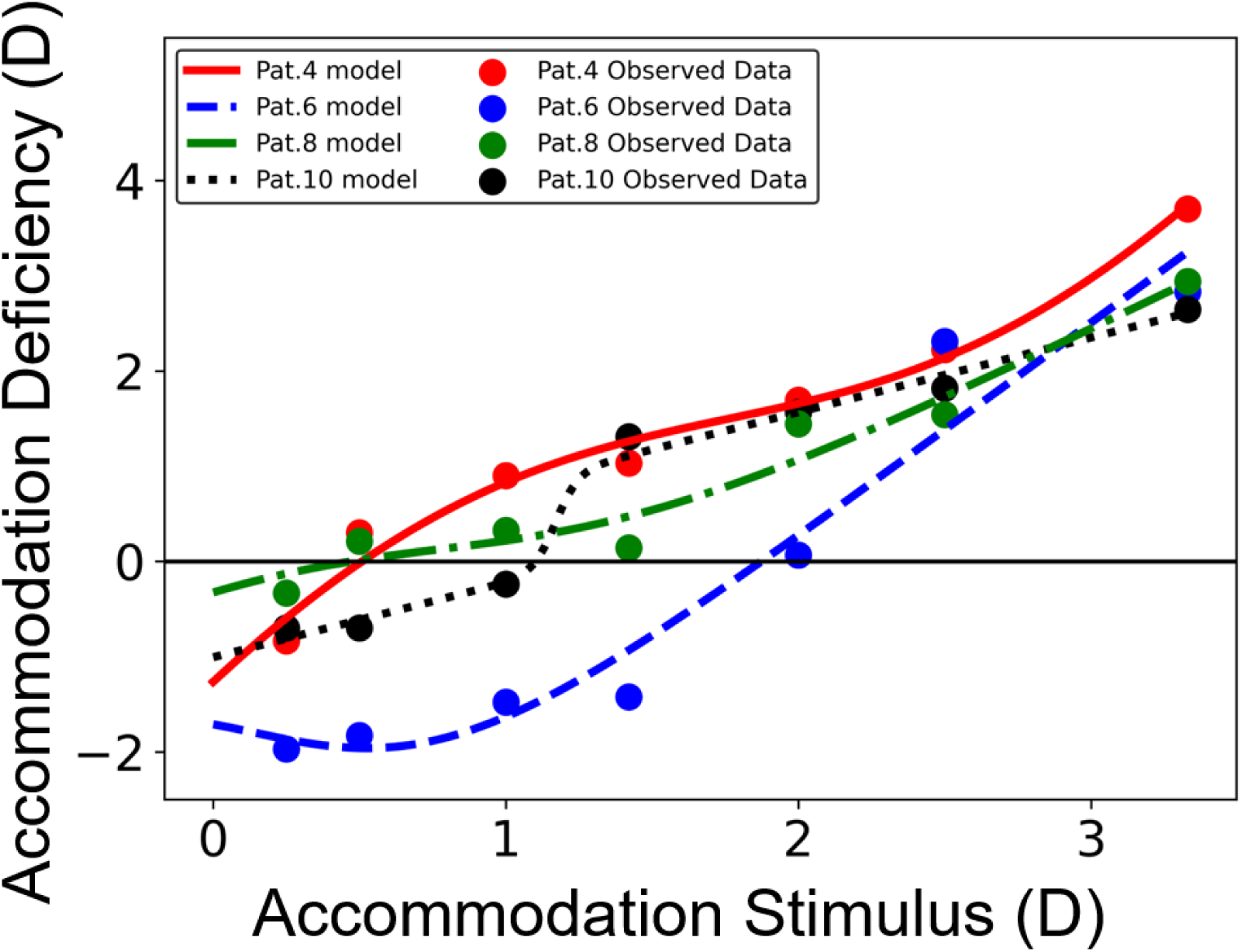
Monocular accommodation deficiency responses and corresponding model fits for different eyes of 4 subjects. The data have been fitted to Equation (1).

## Discussion

Focus-tunable eyewear will likely replace the existing multifocal and progressive eyeglasses in the near future. It is critical that such eyewear be tested on presbyopia patients to evaluate the quality of vision correction which they can provide. However, only one study^11^ has been done so far to evaluate the visual acuity of presbyopes while wearing such devices. To this end, visual acuities of subjects were measured while performing the visual tasks with manually-tunable adjustable-focus eyeglasses. The results of our study show that the corrected visual acuities of all subjects were mostly equal to or better than +0.1 LogMAR (20/25 Snellen equivalent) under all stimulus and illumination conditions. In optometry, visual impairment is defined as visual acuity of worse than +0.3 LogMAR or 20/40 Snellen equivalent^60^. Since the corrected visual acuities of subjects while wearing tunable eyeglasses were better than the visual impairment value, it could be said that the next generation focus-tunable eyewear are indeed a promising replacement to the current eyewear technologies and can provide better vision correction under a host of stimulus and illumination conditions.

A large number of articles written so far have attempted to describe the various mechanisms of presbyopia^20,21,27,61–64^, and they report a steady deterioration in the accommodation response characteristics of the human eye with increasing age and accommodation stimulus^1–7,64–68^. Our study demonstrated that changes in object illumination levels across all object distances also have a significant effect on the corrected visual acuities of presbyopes. The mean visual acuity scores across all distances improved as the chart illumination was increased from 75 lux to 800 lux. These results are consistent with findings of extensive studies demonstrated previously^69–77^. The findings of our study are also consistent with those conducted in these articles^31,33^, where the authors found progressive reduction in the accommodation amplitude with a reduction in target luminance. This intuitively translates to an increase in the AD with reduction in the level of illumination, which can be seen in the results of our study. The improvement in the corrected visual acuity and the AD at higher illumination levels can be explained by the formation of a smaller blur on the retina due to a decrease in pupil diameter and its effects on the depth-of-focus on the human visual system^31^. This article demonstrates the capability of the presented accommodation deficiency model based on the applied accommodation stimulus and the illumination level. 93% of the 30 eyes under examination fit the accommodation deficiency model with root-mean-square error less than 0.5 D. It has been noted in literature that the human eye cannot perceive blur if the defocus error is less than the depth-of-focus of the eye (± 0.45 D – 1.76 D)^78^. Therefore, the analytical model presented in this paper has the potential to accurately describe the accommodation deficiency in presbyopes.

## Conclusions

Presbyopia is an inevitable reality in humans. Adjustable-focus eyewear, which are the future of vision correction technologies, have a crucial role to play in restoring accommodation in presbyopes. However, to perform accurate accommodation correction in such eyewear, a precise understanding of the accommodation deficiency in patients is necessary. Accommodation deficiency data for presbyopic patients under 3 different object illumination conditions has been presented for the first time in literature. This data shows that while accommodation deficiency in humans is a function of the stimulus, it is also strongly dependent on the object illumination. Furthermore, an illumination-dependent model which potentially describes the relation between static accommodation deficiency, accommodation stimulus and object illumination has also been presented in this article. This phenomenological model can be used in accommodation correction devices to partially restore accommodation in presbyopes.

## Data Availability

Data will be made available on the ClinicalTrials.gov registry after the completion of the study.

## References

1. Hamasaki D, Ong J, Marg E. The amplitude of accommodation in presbyopia. Optom Vis Sci. 1956;33(1):3–14. doi:10.1097/00006324-195601000-00002

2. Hofstetter HW. A longitudinal study of amplitude changes in presbyopia. Optom Vis Sci. 1965;42(1):3–8. doi:10.1097/00006324-196501000-00002

3. Ramsdale C, Charman WN. A longitudinal study of the changes in the static accommodation response. Ophthalmic Physiol Opt. 1989;9(3):255–263. doi:10.1111/j.1475-1313.1989.tb00903.x

4. Duane A. Studies in monocular and binocular accommodation with their clinical applications. Am J Ophthalmol. 1922;5(11):865–877. doi:10.1016/S0002-9394(22)90793-7

5. Coates WR. Amplitudes of accommodation in South Africa. Br J Physiol Opt. 1955;12(2).

6. Turner MJ. Observations on the Normal Subjective Amplitude of Accommodation. Br J Physiol Opt. 1958;15(2):70–100.

7. Ayrshire Study Circle: An investigation into accommodation. Br J Physiol Opt. 1964;21:31–35.

8. Hasan N, Banerjee A, Kim H, Mastrangelo CH. Tunable-focus lens for adaptive eyeglasses. Opt Express. 2017;25(2):1221. doi:10.1364/oe.25.001221

9. Hasan N, Karkhanis M, Khan F, Ghosh T, Kim H, Mastrangelo CH. Adaptive optics for autofocusing eyeglasses. In: Optics InfoBase Conference Papers. Vol Part F45-IAO 2017. OSA – The Optical Society; 2017:AM3A.1. doi:10.1364/AIO.2017.AM3A.1

10. Hasan N, Karkhanis M, Ghosh C, et al. Lightweight smart autofocusing eyeglasses. In: Piyawattanametha W, Park Y-H, Zappe H, eds. MOEMS and Miniaturized Systems XVII. Vol 10545. SPIE; 2018:6. doi:10.1117/12.2300737

11. Padmanaban N, Konrad R, Wetzstein G. Autofocals: Evaluating gaze-contingent eyeglasses for presbyopes. Sci Adv. 2019;5(6):eaav6187. doi:10.1126/sciadv.aav6187

12. Jarosz J, Molliex N, Chenon G, Berge B. Adaptive eyeglasses for presbyopia correction: an original variable-focus technology. Opt Express. 2019;27(8):10533. doi:10.1364/oe.27.010533

13. Mastrangelo AS, Karkhanis M, Likhite R, et al. A low-profile digital eye-tracking oculometer for smart eyeglasses. In: Proceedings – 2018 11th International Conference on Human System Interaction, HSI 2018. Institute of Electrical and Electronics Engineers Inc.; 2018:506-512. doi:10.1109/HSI.2018.8431368

14. Ciuffreda KJ. Accommodation and its anomalies. In: Charman WN, ed. Vision and Visual Dysfunction Vol 1: Visual Optics and Instrumentation. Basingstoke: Macmillan; 1991:231-279.

15. Ciuffreda KJ. Accommodation, the pupil and presbyopia. In: Benjamin WJ, ed. Borish’s Clinical Refraction. Philadelphia: Saunders; 1998:77-120.

16. Duane A. Are the current theories of accommodation correct? Am J Ophthalmol. 1925;8(3):196–202. doi:10.1016/S0002-9394(25)90654-X

17. Gullstrand A. Die optische abbildung in heterogen medien die dioptrik det kristallinse des menschen. K Sven Ventenskapsakad Handl.

18. Hess C. Arbeiten aus dem Gebiete der Accommodationslehre. v Graefes Arch f Ophthalmol. 1896;42(1):288.

19. Morgan MW. The ciliary body in accommodation and accommodative-convergence. Optom Vis Sci. 1954;31(5):219–229. doi:10.1097/00006324-195405000-00001

20. Atchison DA. Accommodation and presbyopia. Ophthalmic Physiol Opt. 1995;15(4):255–272. doi:10.1046/j.1475-1313.1995.9500020e.x

21. Charman WN. The eye in focus: Accommodation and presbyopia. Clin Exp Optom. 2008;91(3):207–225. doi:10.1111/j.1444-0938.2008.00256.x

22. von Helmholtz H. Helmholtz’s Treatise on Physiological Optics, Vol 1 (Trans. from the 3rd German Ed.). Optical Society of America; 2011. doi:10.1037/13536-000

23. Fincham EF. The mechanism of accommodation. Br J Opthalmology Monogr Suppl No 8. 1937.

24. Stark L. Presbyopia in light of accommodation. Optom Vis Sci. 1988;65(5):407–416. doi:10.1097/00006324-198805000-00018

25. Donders FC. On the Anomalies of Accommodation and Refraction of the Eye. Translated by Moore WD. New Sydenham Soc. 1984. doi:10.1001/archopht.1986.01050210035014

26. Adler-Grinberg D. Questioning our classical understanding of accommodation and presbyopia. Optom Vis Sci. 1986;63(7):571–580. doi:10.1097/00006324-198607000-00012

27. Pierscionek BK. What we know and understand about presbyopia. Clin Exp Optom. 1993;76(3):83–90. doi:10.1111/j.1444-0938.1993.tb05095.x

28. Semmlow JL, Stark L, Vandepol C, Nguyen A. The Relationship between Ciliary Muscle Contraction and Accommodative Response in the Presbyopic Eye. In: Presbyopia Research. Springer US; 1991:245-253. doi:10.1007/978-1-4757-2131-7_24

29. Fincham EF. The proportion of ciliary muscular force required for accommodation. J Physiol. 1955;128(1):99–112. doi:10.1113/jphysiol.1955.sp005293

30. Shlaer S. The relation between visual acuity and illumination. J Gen Physiol. 1937;21(2):165–188. doi:10.1085/jgp.21.2.165

31. Lara F, Bernal-Molina P, Fernandez-Sanchez V, Lopez-Gil N. Changes in the objective amplitude of accommodation with pupil size. Optom Vis Sci. 2014;91(10):1215–1220. doi:10.1097/0PX.0000000000000383

32. Campbell FW. The minimum quantity of light required to elicit the accommodation reflex in man. J Physiol. 1954;123(2):357–366. doi:10.1113/jphysiol.1954.sp005056

33. Johnson CA. Effects of Luminance and Stimulus Distance on Accommodation and Visual Resolution. J Opt Soc Am. 1976;66(2):138–142. doi:10.1364/J0SA.66.000138

34. Salmon TO, Van De Pol C. Normal-eye Zernike coefficients and root-mean-square wavefront errors. J Cataract Refract SURG. 2006;32. doi:10.1016/j.jcrs.2006.07.022

35. Applegate RA, Donnelly, III WJ, Marsack JD, Koenig DE, Pesudovs K. Three-dimensional relationship between high-order root-mean-square wavefront error, pupil diameter, and aging. J Opt Soc Am A. 2007;24(3):578. doi:10.1364/josaa.24.000578

36. Watson AB, Ahumada AJ. Predicting visual acuity from wavefront aberrations. J Vis. 2008;8(4):1–19. doi:10.1167/8.4.17

37. ClinicalTrials.gov [Internet]. Bethesda (MD): National Library of Medicine (US). 2000 Feb 29 -. Identifier NCT03911596, Smart Autofocusing Eyeglasses; 2019 Apr 11, available from https://clinicaltrials.gov/ct2/show/NCT03911596.

38. Ripps H, Chin NB, Siegel IM, Breinin GM. The Effect of Pupil Size on Accommodation, Convergence, and the AC/A Ratio. Invest Ophthalmol Vis Sci. 1962;1:127–135.

39. Watson AB, Yellott JI. A unified formula for light-adapted pupil size. J Vis. 2012;12(10):12–12. doi:10.1167/12.10.12

40. De Groot SG, Gebhard JW. Pupil size as determined by adapting luminance. J Opt Soc Am. 1952;42(7):492–495. doi:10.1364/JOSA.42.000492

41. Smith G. Relation between spherical refractive error and visual acuity. In: Optometry and Vision Science. Vol 68. Optom Vis Sci; 1991:591-598. doi:10.1097/00006324-199108000-00004

42. Wilcoxon F. Individual Comparisons by Ranking Methods. In: Kotz S, Johnson NL, eds. Breakthroughs in Statistics. Springer Series in Statistics (Perspectives in Statistics). Springer, New York, NY; 1992:196-202. doi:10.1007/978-1-4612-4380-9_16

43. Weale RA. Real light scatter in the human crystalline lens. Graefe’s Arch Clin Exp Ophthalmol. 1986;224(5):463–466. doi:10.1007/BF02173364

44. Aslam TM, Haider D, Murray IJ. Principles of disability glare measurement: an ophthalmological perspective. Acta Ophthalmol Scand. 2007;85(4):354–360. doi:10.1111/j.1600-0420.2006.00860.x

45. Mainster MA, Turner PL. Glare’s causes, consequences, and clinical challenges after a century of ophthalmic study. Am J Ophthalmol. 2012;153(4):587–593. doi:10.1016/j.ajo.2012.01.008

46. Vos JJ. Reflections on glare. Light Res Technol. 2003;35(2):163–176. doi:10.1191/1477153503li083oa

47. van den Berg TJTP, van Rijn LJ, Kaper-Bongers R, et al. Disability glare in the aging eye. Assessment and impact on driving. J Optom. 2009;2(3):112–118. doi:10.3921/joptom.2009.112

48. Spear PD. Neural bases of visual deficits during aging. Vision Res. 1993;33(18):2589–2609. doi:10.1016/0042-6989(93)90218-L

49. Glasser A, Campbell MCW. Presbyopia and the optical changes in the human crystalline lens with age. Vision Res. 1998;38(2):209–229. doi:10.1016/S0042-6989(97)00102-8

50. Curcio CA, Owsley C, Jackson GR. Spare the rods, save the cones in aging and age-related maculopathy. Investig Ophthalmol Vis Sci. 2000;41(8):2015–2018. http://intl.iovs.Org/cgi/content/full/41/8/2015. Accessed July 18, 2020.

51. Glasser A. Presbyopia and aging in the crystalline lens. J Vis. 2010;3(12):22–22. doi:10.1167/3.12.22

52. Wolf E. Glare and Age. Arch Ophthalmol. 1960;64(4):502–514. doi:10.1001/archopht.1960.01840010504005

53. Lasa MSM, Datiles MB, Podgor MJ, Magno B V. Contrast and Glare Sensitivity: Association with the Type and Severity of the Cataract. Ophthalmology. 1992;99(7):1045–1049. doi:10.1016/S0161-6420(92)31852-4

54. Lasa MSM, Podgor MJ, Datiles MB, Caruso RC, Magno B V. Glare sensitivity in early cataracts. Br J Ophthalmol. 1993;77(8):489–491. doi:10.1136/bjo.77.8.489

55. Facchin A, Daini R, Zavagno D. The glare effect test and the impact of age on luminosity thresholds. Front Psychol. 2017;8(JUN). doi:10.3389/fpsyg.2017.01132

56. Levenberg K. A method for the solution of certain non-linear problems in least squares. Q Appl Math. 1944;2(2): 164-168. https://www.ams.org/qam/1944-02-02/S0033-569X-1944-10666-0/. Accessed June 30, 2020.

57. Marquardt DW. An Algorithm for Least-Squares Estimation of Nonlinear Parameters. J Soc Ind Appl Math. 1963;11(2):431–441. doi:10.1137/0111030

58. Shieh G. Improved Shrinkage Estimation of Squared Multiple Correlation Coefficient and Squared Cross-Validity Coefficient. Organ Res Methods. 2008;11(2):387–407. doi:10.1177/1094428106292901

59. Theil H, Cramer JS. Economic Forecasts and Policy. North-Holland Publishing Company; 1961.

60. Chia EM, Wang JJ, Rochtchina E, Smith W, Cumming RR, Mitchell P. Impact of Bilateral Visual Impairment on Health-Related Quality of Life: The Blue Mountains Eye Study. Investig Ophthalmol Vis Sci. 2004;45(1):71–76. doi:10.1167/iovs.03-0661

61. Weale R. Presbyopia toward the end of the 20th century. Surv Ophthalmol. 1989;34(1):15–30. doi:10.1016/0039-6257(89)90126-4

62. Croft MA, Glasser A, Kaufman PL. Accommodation and presbyopia. Int Ophthalmol Clin. 2001;41(2):33–46. doi:10.1097/00004397-200104000-00005

63. Gilmartin B. The Aetiology of Presbyopia: A Summary of the Role of Lenticular and Extralenticular Structures. Ophthalmic Physiol Opt. 1995;15(5):431–437.

64. Mordi JA, Ciuffreda KJ. Static aspects of accommodation: Age and presbyopia. Vision Res. 1998;38(11):1643–1653. doi:10.1016/S0042-6989(97)00336-2

65. Kalsi M, Heron G, Charman WN. Changes in the static accommodation response with age. Ophthalmic Physiol Opt. 2001;21(1):77–84. doi:10.1046/j.1475-1313.2001.00546.x

66. Fitch RC. Procedural effects on the manifest human amplitude to accommodation. Optom Vis Sci. 1971;48(11):918–925. doi:10.1097/00006324-197111000-00004

67. Somers WW, Ford CA. Effect of relative distance magnification on the monocular amplitude of accommodation. Optometery Vis Sci. 1983;60:920–924.

68. Atchison DA, Capper EJ, McCabe KL. Critical subjective measurement of amplitude of accommodation. Optom Vis Sci. 1994;71(11):699–706. doi:10.1097/00006324-199411000-00005

69. Tidbury LP, Czanner G, Newsham D. Fiat Lux: the effect of illuminance on acuity testing. Graefe’s Arch Clin Exp Ophthalmol. 2016;254(6):1091–1097. doi:10.1007/s00417-016-3329-7

70. Wozniak HP, Kelly M, Glover S, Moss ND. The effect of room illumination on visual acuity measurement. Aust Orthopt J. 1999;(34):3-8.

71. Sheedy JE, Bailey IL, Raasch TW. Visual acuity and chart luminance. Optom Vis Sci. 1984;61(9):595–600. doi:10.1097/00006324-198409000-00010

72. Rabin J. Luminance effects on visual acuity and small letter contrast sensitivity. Optom Vis Sci. 1994;71(11):685–688. doi:10.1097/00006324-199411000-00003

73. Lee EM, Feis AE, Clark A. Effect of room illumination in computerized visual acuity (using smart system II). Optometry. 2009;80(6):316. doi:10.1016/j.optm.2009.04.072

74. Johnson CA, Casson EJ. Effects of luminance, contrast, and blur on visual acuity. Optom Vis Sci. 1995;72(12):864–869. doi:10.1097/00006324-199512000-00004

75. Simpson TL, Barbeito R, Bedell HE. The Effect of Optical Blur on Visual Acuity for Targets of Different Luminances. Ophthalmic Physiol Opt. 1986;6(3):279–281. doi:10.1111/j.1475-1313.1986.tb00716.x

76. Haymes SA, Lee J. Effects of task lighting on visual function in age-related macular degeneration. Ophthalmic Physiol Opt. 2006;26(2):169–179. doi:10.1111/j.1475-1313.2006.00367.x

77. Baker PA, Raos AS, Thompson JMD, Jacobs RJ. Visual acuity during direct laryngoscopy at different illuminance levels. Anesth Analg. 2013;116(2):343–350. doi:10.1213/ANE.0b013e318273f397

78. Wang B, Ciuffreda KJ. Depth-of-focus of the human eye: Theory and clinical implications. Surv Ophthalmol. 2006;51(1):75–85. doi:10.1016/j.survophthal.2005.11.003

